# Postoperative Determination of Directional Deep Brain Stimulation (DBS) Lead Orientation Using Rotational Fluoroscopy: Interobserver Agreement and Comparison with CT-Based Software

**DOI:** 10.1101/2025.08.24.25334315

**Authors:** Paresh K Doshi, Chandresh Karnavat, Raj V Agarbattiwala

**Author notes:** Corresponding Author: Prof. Paresh K Doshi Address: Jaslok Hospital and Research Center, 15, G Deshmukh Marg, Pedder Road, Mumbai-40026.

## Abstract

**Background:** Directional deep-brain stimulation (DBS) requires knowledge of lead orientation to maximize benefit and minimize side effects. While CT-based software is widely used, its accuracy may decrease with the use of oblique leads. Rotational fluoroscopy using the Iron Sight method offers an alternative; however, its reliability in clinical practice has not been fully established.

**Objective:** To assess the reliability of the Iron Sight method for postoperative lead orientation and compare it with CT-based software (Brainlab Elements™).

**Methods:** We prospectively analyzed 70 directional leads implanted at our center (2022–2024). Orientation was measured on postoperative rotational fluoroscopy by two independent observers and compared with postoperative CT analyzed in Brainlab Elements™. The agreement was evaluated using established reliability statistics.

**Results:** Observers using the Iron Sight method produced highly consistent measurements, differing on average by <1° and rarely more than ±7°. Compared with CT-based analysis, the Iron Sight method showed good agreement, with differences typically within ±15°. These margins correspond to <1.5 mm displacement around the lead — well within clinically acceptable limits for programming.

**Conclusion:** Rotational fluoroscopy, combined with the Iron Sight method, provides clinicians with a reliable and reproducible tool to confirm lead orientation. It may be especially valuable when CT-based software is unavailable, inconclusive, or requires corroboration.

## Introduction

Deep brain stimulation (DBS) is a recognized treatment for movement disorders like Parkinson’s disease, tremors, and dystonia, as well as drug-resistant epilepsy and obsessive-compulsive disorder, the latter still being experimental (1–4). The clinical scope of DBS continues to expand, with ongoing research exploring its role in psychiatric and neurodegenerative conditions including major depression, addiction, Tourette syndrome, Alzheimer’s disease, and schizophrenia.

Despite decades of clinical use, the technology underlying DBS systems remained unchanged until the recent introduction of directional leads. Traditional cylindrical DBS electrodes produce approximately spherical volumes of tissue activation, often leading to the unintended stimulation of adjacent structures. The potential benefits of directing stimulation between contacts or in specific directions have long been anticipated (5). Directional DBS leads to feature-segmented electrodes that allow clinicians to steer the electrical field, improve therapeutic precision, expand the therapeutic window, and reduce stimulation-induced side effects (6,7). This steering is particularly beneficial in targeting areas such as the subthalamic nucleus (STN), where directional stimulation can avoid spreading to the internal capsule (IC) while enhancing the coverage of functionally relevant pathways.

Accurate knowledge of the final orientation of the segmented contacts is crucial for individualized stimulation planning (8–11). Although electrodes are inserted with the intended orientation (typically anterior), intraoperative lead rotation and torsion can alter the final position. Despite the directional markers in existing commercial models, accurately determining the orientation of the lead continues to pose difficulties (12). Consequently, postoperative imaging is essential to confirm lead orientation. Among various imaging techniques, postoperative computed tomography (CT) combined with preoperative magnetic resonance imaging (MRI) and interpretation using commercial software (e.g., Brainlab Elements™) is a common practice (13). However, CT-based methods may lose accuracy when leads are positioned at oblique angles (>40°) (14).

Techniques such as the “Iron Sight” method, which exploits the radiographic appearance of segmented contacts to infer orientation, have demonstrated high accuracy in phantom models. Reinacher et al. reported a limit of agreement (LoA) of ±2.44° and an intraclass correlation coefficient (ICC) of 0.9998 using this method (12). However, its inter-rater reliability in real-world clinical scenarios has not been sufficiently studied. Furthermore, the agreement between fluoroscopy-based estimates and software-derived CT orientations has not yet been systematically quantified.

## Aim and Objectives

### Aim

To evaluate the inter-rater agreement in determining directional DBS lead orientation using the Iron Sight method on postoperative 3D rotational fluoroscopy and to compare these observer-derived estimates with orientation measurements from Brainlab Elements™ software based on postoperative CT imaging.

### Objectives

- To assess interobserver agreement between a neurosurgeon and a radiologist in determining directional DBS lead orientation using the iron-sight method on postoperative 3D rotational fluoroscopy.
- To compare orientation measurements obtained via the Iron Sight method with those derived from the Brainlab Elements™ software using postoperative CT.
- To assess the association of lead trajectory angles with measurement errors by the above methods
- The reliability and statistical uncertainty of the assessments were quantified using circular statistics and intraclass correlation.

## Methods

Reporting in the article follows the Guidelines for Reporting Reliability and Agreement Studies (GRRAS) to ensure the full disclosure of design and statistical decisions. This prospective exploratory study was conducted at a high-volume tertiary care center between May 2022 and December 2024.

### Participants

All consecutive patients who underwent DBS surgery with directional leads (Vercise Cartesia™, Boston Scientific) during the study period were screened for inclusion. The inclusion criteria were as follows.

- Implantation of directional DBS leads (Vercise Cartesia™, Boston Scientific) for movement disorders.
- The participant (or legally authorized representative) must have given consent.

Patients with incomplete imaging, significant motion artifacts, or postoperative complications affecting the lead visualization were excluded.

The study was conducted in accordance with the Declaration of Helsinki and was approved by the Institutional Ethics Committee at our center (approval number EC/1114/2022, dated April 29, 2022). Participants were recruited from among patients who underwent DBS surgery with directional leads (Vercise Cartesia™, Boston Scientific) between May 1, 2022, and December 31, 2024. All patients and caregivers received both verbal and written explanations of the study, and written consent was obtained from each participant (or their representative) for participation and secondary analysis of the de-identified data. Both CT and fluoroscopy were performed during the postoperative period after admission. All surgeries were conducted under the Stereotactic and Functional Neurosurgery department at our center by a single neurosurgeon (PD) according to the standard protocol (15). CT scans were performed in the radiology department, and rotational fluoroscopy was performed in the Cath lab (see Table A of the Supplementary Table for the parameters).

### Sample Size

Sample size estimation for comparing rater reliability or agreement has not been thoroughly explored. Fleiss proposed a rule of thumb that recommends 15–20 samples for reliability studies (16). In the case of a continuous outcome, increasing the sample size beyond 12 per group did not significantly affect the confidence interval. Reinacher et al. calculated a 95% confidence interval of ±2.44 ° in their study of phantom models. Using Zou’s formula for ICC precision with 95% CI width ≤ 0.1, α = 0.05, ICC = 0.85, and 80% assurance, the required sample size was 57 leads for two raters (17). We chose an expected ICC of 0.85 and a 95% CI width of ≤ 0.1 because ICC ≥ 0.75 suggests good reliability. Our study included 70 leads that met this threshold (18). However, we included 70 leads to account for real-world variability in human data and to ensure robustness in statistical comparisons.

### Postoperative Imaging and Orientation Measurement

#### Rotational Fluoroscopy (Iron Sight Method)

Within 48 hours postoperatively, all patients were brought to Cathlab for 3D rotational fluoroscopy. The acquisition parameters are listed in the Supplementary Table. Images were reconstructed using multiple projections and independently reviewed by two raters.

- A neurosurgeon (RA) experienced in DBS surgery and programming
- A radiologist (CK) trained in neuroimaging

The observers performed measurements independently on anonymized files and had no access to each other’s results. Using the Iron Sight method, each observer independently estimated the rotational orientation of each lead relative to the anterior direction, expressed in degrees (–180° to +180°) (Supplementary Data).

#### Determination of lead orientation on CT scans

Postoperative CT scans were uploaded into Brainlab Elements™ (Lead Localization module). The acquisition parameters are listed in Table A of the Supplementary Tables. The system automatically detected the lead axis and segmented the contact positions based on CT artifact patterns (as described by Hellerbach et al., 2018 (19)), which were then visually confirmed. The lead model was set to Boston Scientific Vercise Cartesia™. The output orientation is recorded as the reference value for each lead. Given that Brainlab Elements™ itself has known limitations at oblique angles, this method comparison should be interpreted as assessing agreement between the two techniques rather than validating one against a true gold standard. No phantom-based ground truth data were available in this clinical setting.

### Statistical Analysis

All statistical analyses were performed using MATLAB (MathWorks, Natick, MA, USA) and the Circular Statistics Toolbox (20). Statistical significance was set *P-value < 0.05*.

#### Interobserver Agreement

To assess interobserver agreement using the Iron Sight method, orientation angles were treated as circular variables ranging from –180° to +180°. The circular difference, or shortest angular distance, was calculated for each lead between the two raters’ measurements. Circular variance was computed to quantify the dispersion of the ratings, and the circular correlation coefficient (21) was used to assess the strength of association between the estimates of the neurosurgeon and radiologist. Because angular dispersion was low, we used ICC (2,1), as defined by Shrout and Fleiss (1979) (22), which is appropriate for two-way random effects models assessing absolute agreement between two observers. This ICC formulation accounts for subject variability, rater variability, and residual errors.

A Bland–Altman analysis (23,24) adapted for circular data (Online Appendix) was conducted to further explore the agreement. This included calculating the mean angular difference (i.e., circular bias) and standard deviation of the differences. The 95% limits of agreement (LoA) were computed as the bias ± 1.96 of the standard deviation. To express the uncertainty in these estimates, 95% confidence intervals for both the bias and limits of agreement were calculated. Finally, the Preiss-Fisher procedure was applied to test whether the observed agreement could have occurred by chance (25).

#### Method Comparison (Ironsight method vs Brainlab™)

For method comparison between the iron-sight approach and Brainlab Elements™, the Watson–Williams test was used to compare the circular means derived from the observers’ average ratings with the angles obtained from the Elements™ software(26). The Intraclass Correlation Coefficient (ICC (3,1)) was calculated to quantify the consistency between the two methods, using a two-way mixed-effects model. Agreement between the iron-sight method and Brainlab™-derived orientation angles was also visualized using a Bland–Altman plot. The Preiss-Fisher test was again applied to confirm whether the observed concordance between the methods exceeded chance expectations. Before the parametric circular tests, assumptions were assessed and met, including angular homogeneity and concentration.

#### Regression Analysis

We performed two univariate linear regression analyses to investigate whether anatomical trajectory angles influenced the angular measurement differences. Linear regression of the coronal, sagittal, polar, and azimuthal angles (as explained by Matias et al. (27)) against the angular difference between the two observers’ measurements on rotational fluoroscopy (i.e., interobserver error). A separate regression of these angles against the angular differences between rotational fluoroscopy and Brainlab Elements™ (i.e., method discrepancy).

For each regression model, beta coefficients and the corresponding p-values were calculated to assess the strength and significance of the association between the variables. Owing to multiple comparisons, we adjusted the p-values (P_adj_) for the false discovery rate using the Benjamini-Hochberg method (28). Statistical significance was set at *P* <0.05.

## Results

### Study Sample

Thirty-five patients were included in the final analysis, yielding 70 directional DBS leads for inter-observer agreement analysis. For the method comparison analysis, five leads were excluded because the Brainlab Elements™ software failed to compute the lead orientation, resulting in 67 leads being analyzed. An overview of the data, including circular means, ranges, and standard deviations for all measurements, is presented in Table 1.

**Table 1.** Descriptive Statistics of Directional DBS Lead Orientations Estimated by Observers and Brainlab Elements™.

|  |  |  |
| --- | --- | --- |
| Measurements | circular mean | Circular Standard |

|  | (angular range) (in degrees) | Deviation (in degrees) |
| --- | --- | --- |
| Angles measured by observer 1 (Neurosurgeon) (n = 70) | 5.55° (-64° to 42°) | 16.89° |
| Angles measured by observer 2 (Radiologist) (n = 70) | 4.89° (-67° to 39°) | 16.36° |
| Angles measured by Brainlab Element™ (n = 67) | 8.27° (-29° to 46°) | 13.51° |
| Trajectories Coronal Angle (n = 70) | 24.24° (15.64° to 36.45°) | 4.33° |
| Trajectories Sagittal Angle (n = 70) | 25.84° (11.31° to 41.15°) | 6.38° |
| Trajectories Axial Angle (n = 70) | 43.38° (21.21° to 70.45°) | 10.09° |
| Trajectories Azimuth Angle (n = 70) | 33.95° (24.48° to 46.74°) | 4.57° |

Descriptive summary of lead orientation measurements from postoperative rotational fluoroscopy and CT in 70 directional leads implanted at our center (2022–2024). Circular means, ranges, and standard deviations are provided for observer estimates using the iron-sight method and Brainlab Elements™ software.

#### Interobserver Agreement: Iron Sight Method on Rotational Fluoroscopy

The orientations estimated by the two observers were visualized using polar histograms (Figure 1A-D). The Rayleigh test for uniformity demonstrated significant data clustering (*P < 0.05*), indicating that the angles were not randomly distributed around the circle, but were concentrated around a common direction.

**Figure 1.**
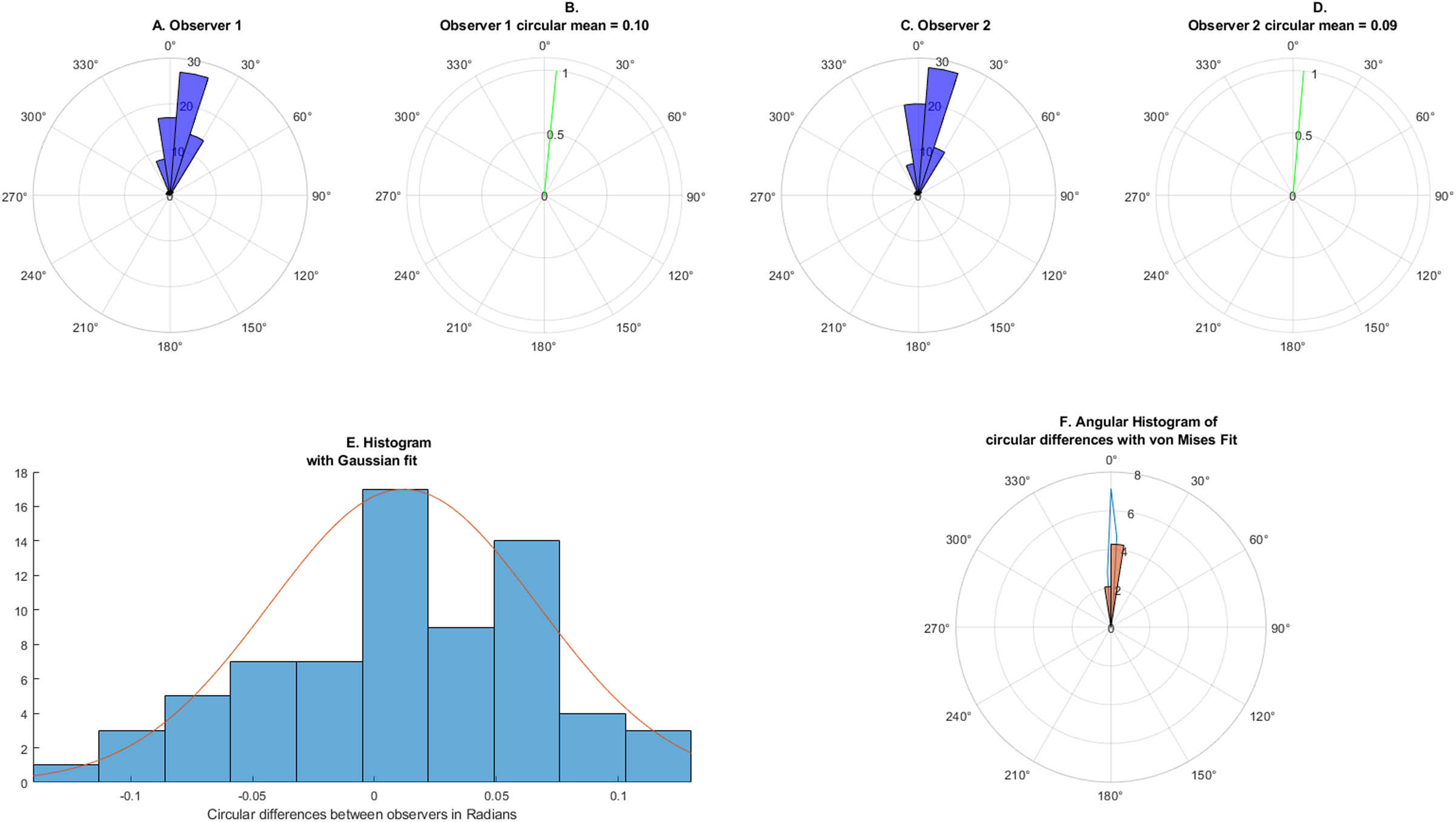
Polar histograms of directional DBS lead orientations measured with the Iron Sight method from postoperative rotational fluoroscopy at Jaslok Hospital, Mumbai (2022-2024). Panel A-D: Orientation estimates from each observer (n=70 leads). Panel E: Distribution of angular differences between observers, showing a symmetric pattern consistent with a Gaussian distribution. Panel F: Polar histogram of observer differences compared with the von Mises distribution. Clinically, differences between observers were minimal, with 95% of measurements within – 5.49° to +6.85° (approximately ±7°).

The mean resultant length of the circular distance between the two observers was 0.9985 with a circular variance of 0.0015, indicating highly consistent measurements. The circular correlation coefficient was 0.9822, confirming a strong directional agreement. A Circular Bland–Altman analysis (figure 2A) revealed a mean bias of 0.68° (95% CI: –0.0678° to 1.4335°). The upper limit of agreement (LoA) was 6.85° (95% CI: 5.55° to 8.15°) and the lower LoA was –5.49° (95% CI: – 6.79° to 4.19°). There was no evidence of proportional bias (*P > 0.05*), suggesting that the measurement errors did not change with angle. The Preiss–Fisher test showed that the standard deviation of differences between two observer measurements yielded a Z-value of –15.40 (Figure 2B), indicating that the observed agreement was significantly greater than would be expected by chance. The Intraclass Correlation Coefficient (ICC (2,1)) was 0.983 (95% CI: 0.973–0.989), indicating excellent inter-observer reliability (Figure 2C and Table B of the Supplementary Tables for calculation parameters).

**Figure 2.**
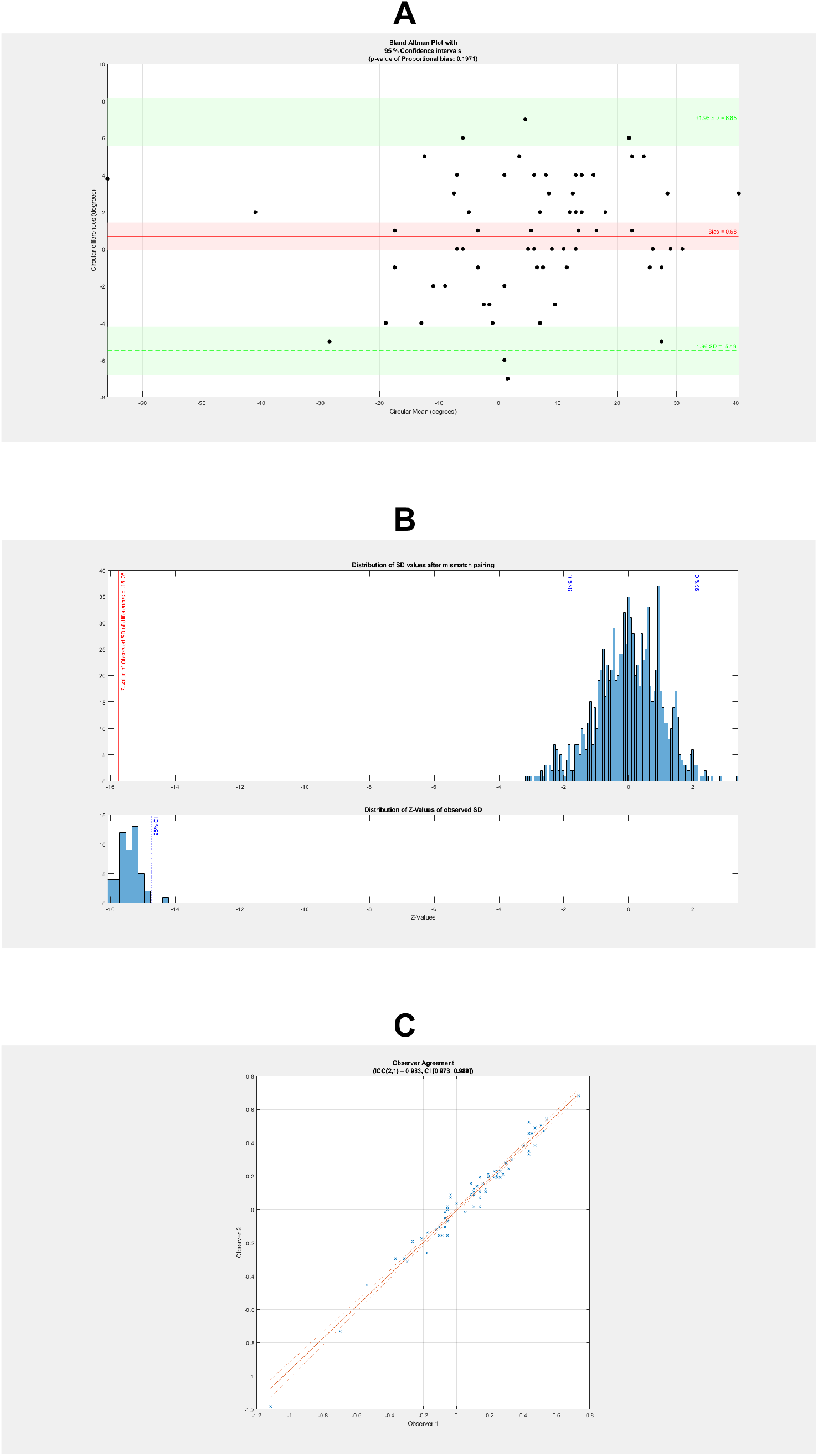
Agreement analysis between neurosurgeon and radiologist estimates of DBS lead orientation using the Iron Sight method on postoperative fluoroscopy (2022-2024). Panel A: Bland–Altman plot showing a mean difference of 0.68° with 95% limits of agreement from –5.49° to +6.85°. Panel B: Preiss–Fisher test confirming agreement was highly unlikely due to chance. Panel C: Intraclass correlation coefficient (ICC(2,1) = 0.983, 95% CI 0.973–0.989) demonstrating excellent interobserver reliability. In practice, this means two trained observers almost always agreed within ∼7° when using rotational fluoroscopy.

Figure 1E shows that the histogram of the circular distances between observers exhibited a symmetric distribution, approximating a Gaussian shape. Similarly, the polar histogram of angular differences was visually compared with the von Mises distribution (Figure 1F) and showed good conformity.

Univariate linear regression analysis revealed that the coronal angle (β = 0.1803, *P-value = 0.0367*; *adjusted P_adj_ = 0.073*) and polar angle (β = 0.0792, *P-value = 0.0320*; *adjusted P_adj_ = 0.073*) were associated with greater interobserver differences. Note that after controlling for the false discovery rate using the Benjamini–Hochberg procedure, these associations did not remain statistically significant at the threshold of 0.05. The sagittal angle (β = –0.0588, *P-value = 0.3212*; *adjusted P_adj_ = 0.428*) and azimuthal angle (β = 0.0364, *P-value = 0.6618*; *adjusted P_adj_ = 0.662*) showed no significant association with interobserver variation.

### Method Comparison: Iron Sight Method vs. Brainlab Elements™

Of the 70 leads, 67 were eligible for method comparison, as the software failed to estimate the orientation in the three leads. Orientation estimates from both methods are displayed using polar histograms (figure 3A-D), including circular means. The Rayleigh test again showed significant nonuniformity (*P < 0.05*), indicating that the data points were directionally concentrated.

**Figure 3.**
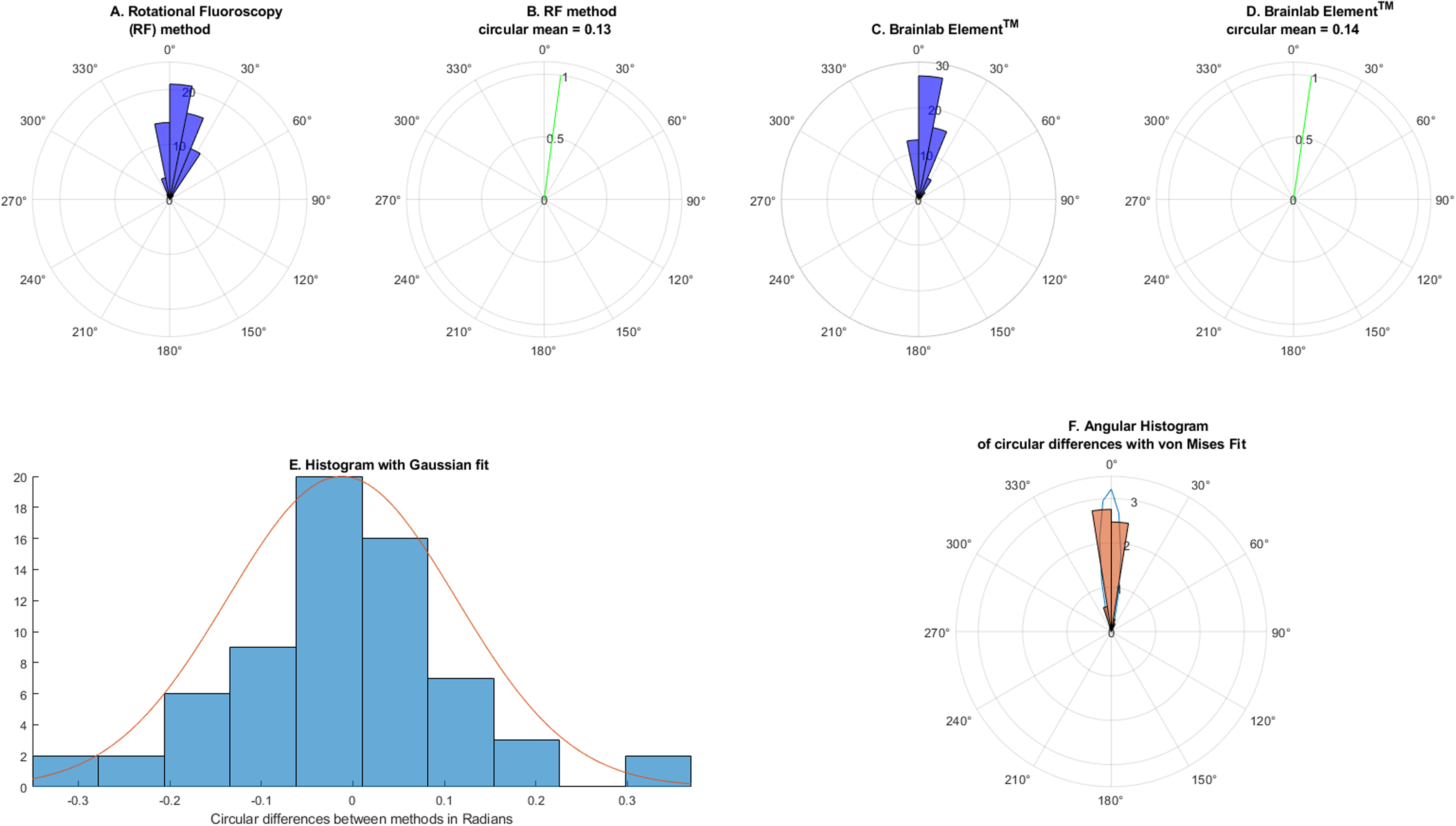
Comparison of the Iron Sight method and Brainlab Elements™ CT-based software in patients (Jaslok Hospital, 2022-2024). Panel A-D: Orientation estimates from both methods for 67 leads. Panel E: Distribution of angular differences approximating a Gaussian shape. Panel F: Polar histogram of differences compared with the von Mises distribution. On average, the two methods differed by less than 1°, with 95% of measurements within –14.86° to +13.48° (about ±15°).

The mean resultant length of the circular distance between the iron-sight method and the Brainlab™-derived orientation was 0.9922, with a circular variance of 0.0078. The circular correlation coefficient between the two methods was 0.8635. Bland–Altman analysis (figure 4A) revealed a mean bias of –0.69° (95% CI: – 2.4839° to 1.0993°), with an upper LoA of 13.48° (95% CI: 10.37° to 16.58°) and a lower LoA of –14.86° (95% CI: –17.97° to –11.76°). No significant proportional bias was observed (*P > 0.05*), and the Preiss–Fisher Z-value was –10.83 (Figure 4B), again indicating that agreement between methods was not attributable to chance. The Intraclass Correlation Coefficient (ICC (3,1)) for the agreement between the Iron Sight method and Brainlab Elements™ was 0.864 (95% CI: 0.786–0.915) (Figure 4C and Table C of the Supplementary Tables for calculation parameters), suggesting good consistency between the two approaches.

**Figure 4.**
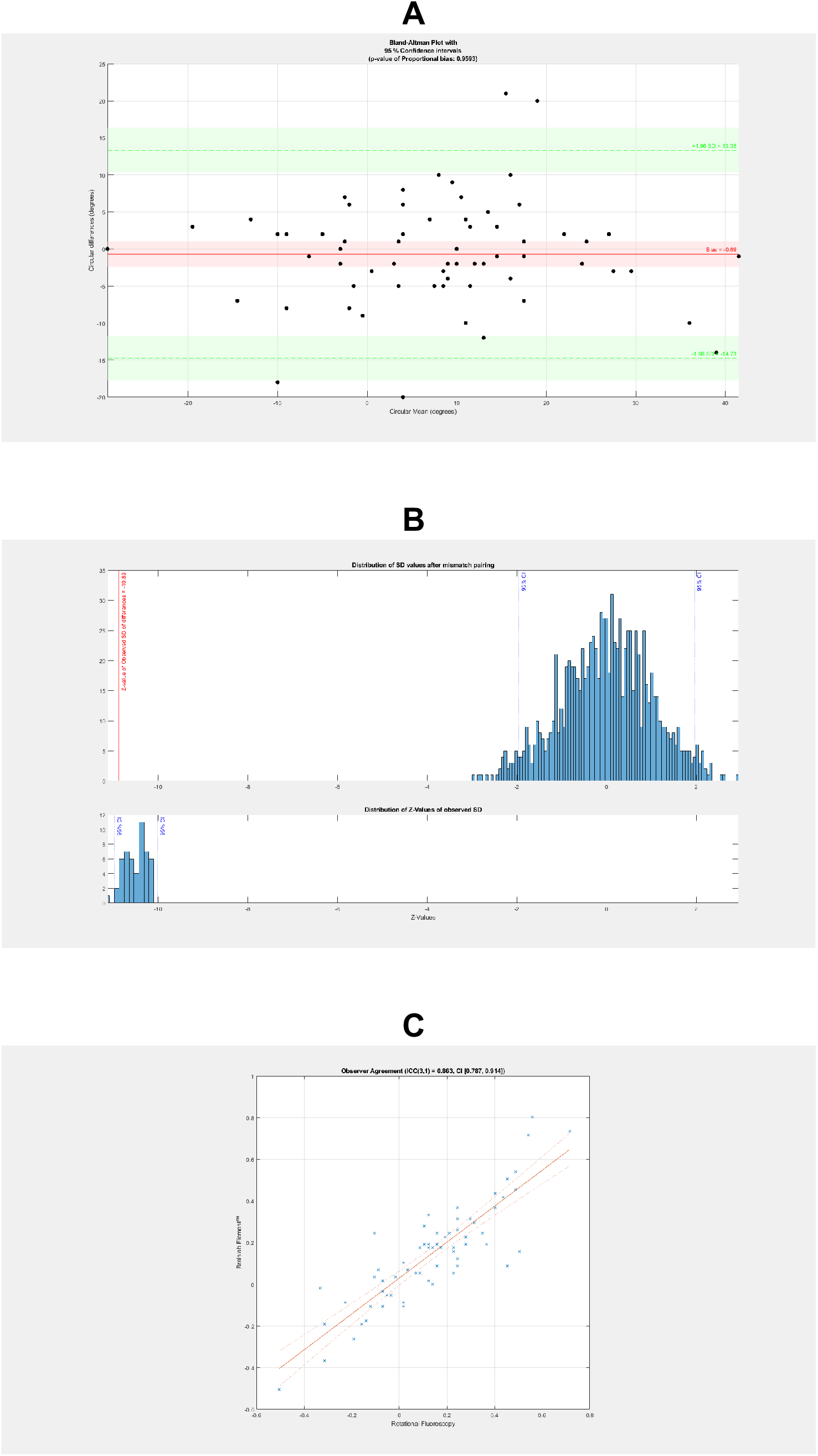
Agreement between rotational fluoroscopy (Iron Sight method) and Brainlab Elements™ software in determining orientation of directional DBS leads (67 leads, Jaslok Hospital, Mumbai, 2022-2024). Panel A: Bland–Altman plot showing a mean bias of –0.69° and 95% limits of agreement from –14.86° to +13.48°. Panel B: Preiss–Fisher test confirming concordance was not due to chance. Panel C: Intraclass correlation coefficient (ICC(3,1) = 0.864, 95% CI 0.786–0.915) indicating good agreement between the two approaches. For clinicians, this means that both fluoroscopy and CT-based methods are broadly consistent, with most measurements agreeing within ∼15° — a margin corresponding to less than 1.5 mm around the electrode circumference.

Figure 3E shows that the histogram of the circular distances between the methods approximates a Gaussian distribution. Similarly, the polar histogram of angular differences showed comparable clustering and was visually comparable to the von Mises distribution (Figure 3F). The Watson–Williams test yielded a *P-value* of *0.7905 (>0.05)*, indicating no statistically significant difference in the circular mean orientations between the two methods.

None of the lead trajectory angles demonstrated a statistically significant relationship with the angular difference between the fluoroscopy-derived and Brainlab-derived orientations. The beta coefficients for the coronal, sagittal, polar, and azimuthal angles were 0.2546 (*P-value = 0.2374, adjusted P_adj_ = 0.7046*), 0.0315 (*P-value = 0.8073, adjusted P_adj_ = 0.8073*), 0.0217 (*P-value = 0.7976, adjusted P_adj_ = 0.8073*), and 0.1688 (*P-value = 0.3523, adjusted P_adj_ = 0.7046*), respectively.

## Discussion

In this study, we evaluated whether rotational fluoroscopy using the Iron Sight method can serve as a reliable tool for determining the orientation of directional deep brain stimulation (DBS) leads. We also compared these results with postoperative CT-based analysis using Brainlab Elements™. To our knowledge, this is among the first clinical studies to assess the interobserver agreement of the Iron Sight method in a real-world patient population using circular statistical methods and to systematically compare these estimates with those of a commercial CT-based platform (Brainlab Elements™).

Our results demonstrate that two independent observers, a neurosurgeon and a radiologist, achieved excellent agreement when using the Iron Sight method. On average, their measurements differed by less than 1°, and in almost all cases the difference was within ±7°. This indicates that, when properly trained, clinicians can consistently determine lead orientation from rotational fluoroscopy images. This real-world evidence supports previous phantom-based work by Reinacher et al., who reported a similar accuracy of the Iron Sight method with a ±2.44° limit of agreement and ICC of 0.9998 in controlled settings (12).

When comparing the Iron Sight method to Brainlab Elements™, the two approaches showed good overall agreement. The mean difference between methods was less than 1° (–0.69°), and in most cases, results fell within ±15°. Although this range is wider than the differences observed between observers, it corresponds to less than 1.5 mm displacement around the electrode circumference — a margin considered clinically acceptable for programming.

The broader limits of agreement likely reflect intrinsic differences between the two methods. The Iron Sight technique relies on direct visualization of segmented contacts, while the CT-based approach depends on artifact symmetry. The latter is known to lose accuracy when electrodes are angled steeply (>40° relative to the scanner axis) (14,29). Nevertheless, under typical implant conditions, both methods produced broadly comparable estimates, and the Watson–Williams test confirmed no statistically significant difference in average orientation values.

We also examined whether lead trajectory angles (coronal, sagittal, polar, and azimuthal) contributed to measurement variability. There was a suggestion that more lateral or off-vertical leads (coronal and polar angles) increased variability between observers. However, these associations did not remain significant after correction for the false discovery rate using the Benjamini-Hochberg method (28). Importantly, none of the trajectory angles meaningfully affected the agreement between fluoroscopy- and CT-based estimates. This suggests that both methods are robust across a wide range of implant trajectories in routine practice.

Orientation accuracy matters because even modest misalignments can influence therapeutic outcomes. Computational models have shown that a 45° misorientation can substantially reduce activation of target neurons or increase the risk of stimulating adjacent structures such as the internal capsule (30). In this context, our observed differences — within ±7° for observers and ±15° between methods — fall well below clinically concerning thresholds. These results reassure clinicians that both rotational fluoroscopy and CT-based software provide sufficiently precise orientation data for safe and effective programming.

Rotational fluoroscopy has additional practical value. It is widely available, can be performed quickly after surgery, and does not require specialized software. It may therefore be especially useful in cases where CT-based analysis is inconclusive, unavailable, or requires corroboration.

### Limitations

This study had several limitations. First, although this was a prospective study, it was conducted at a single institution, with all surgeries performed by the same neurosurgeon using a single imaging system and a single lead model (Boston Scientific Vercise Cartesia™). The generalizability to other hardware platforms or vendors may be limited. Second, the Iron Sight method, although reproducible in our hands, may depend on the observer’s experience and fluoroscopic image quality; training and standardization could impact its reliability for broader use. Furthermore, we compared the Iron Sight method with Brainlab Elements™, which itself may not represent a perfect ground truth. Our reported agreement therefore reflects method concordance rather than absolute accuracy.

## Conclusion

In conclusion, the iron-sight method, when applied to postoperative rotational fluoroscopy, offers excellent interobserver reliability for determining directional DBS lead orientation and demonstrates good agreement with Brainlab Elements™ CT-based estimations. These findings support the clinical utility of rotational fluoroscopy as a reliable imaging modality for orientation assessment, particularly in settings in which software-based methods are unavailable, fail, or require corroboration. Future work should examine reproducibility across different institutions, hardware platforms, and surgical teams. Studies linking orientation accuracy to clinical outcomes such as programming thresholds, therapeutic windows, and stimulation-related side effects will help determine whether small differences in orientation measurement translate into meaningful differences for patients.

## Supporting information

Anonymized Data.xlsx

GRRAS

## Data Availability

All data produced in the present work are contained in the supplementary material.

## Acknowledgements

We thank Jaslok Hospital and Research Institute for their generous support in funding and providing space for our intervention activities. We thank Dr. Manish Baldia and Dr. Neha Rai for their assistance in setting up the initial protocol and coordinating its implementation.

## Autor Roles

1. Research project: A. Conception, B. Organization, C. Execution;
2. Statistical Analysis: A. Design, B. Execution, C. Review and Critique;
3. Manuscript Preparation: A. Writing of the first draft, B. Review and Critique;

Doshi P: 1A, 1B, 2A, 2C, 3B

Karnavat C: 1C, 2B, 3B

Agarbattiwala R: 1B, 1C, 2A, 2B, 2C, 3A, 3B

## Financial Disclosures for the previous 12 months

The authors declare that there are no additional disclosures to report. Ethical Compliance Statement:

- The study was conducted in accordance with the Declaration of Helsinki and was approved by the Institutional Ethics Committee at our center (approval number EC/1114/2022, dated April 29, 2022).
- Written consents were obtained from all the patients.
- We confirm that we have read the Journal’s position on issues involved in ethical publication and affirm that this work is consistent with those guidelines.

## Data Sharing Statement

The data supporting this study’s findings were anonymized and are available in the Supplementary Data.

## Supplementary Data

|  |  |
| --- | --- |
| Appendix | This document describes the mathematical formulation to calculate bias, limits of agreement (LoA), and confidence intervals in a Bland-Altman analysis for circular data |
| Supplementary Tables | Table A lists imaging acquisition parameters, Table B outlines the ICC calculation parameters for interobserver agreement in rotational fluoroscopy, Table C details the parameters for assessing agreement between the methods. |
| Supplementary Data | Angle measurement conventions and approaches by the iron-sight method |
| Anonymized data | Angular data used for inter-rater agreement for the iron-sight method, comparison between the two methods, and regression analysis |

