## Supplementary material for "Postoperative Determination of Directional Deep Brain Stimulation (DBS) Lead Orientation Using Rotational Fluoroscopy: Interobserver Agreement and Comparison with CT-Based Software": GRRAS

Checklist of the Guidelines for Reporting Reliability and Agreement Studies (GRRAS).

| Section | Guidelines | Pages |
| --- | --- | --- |
| TITLE AND ABSTRACT | Identify in title or abstract that interrater/intrarater reliability or agreement was investigated. | 1 - 5 |
| INTRODUCTION | Name and describe the diagnostic or measurement device of interest explicitly. | 5 – 7 |
|  | Describe what is already known about reliability and agreement and provide a rationale for the study (if applicable). | 5 - 7 |
| AIM AND OBJECTIVES | Specify the subject population of interest. | 7 |
|  | Specify the rater population of interest (if applicable). | 7 |
| METHODS | Explain how the sample size was chosen. State the determined number of raters, subjects/objects, and replicate observations. | 9 |
|  | Describe the sampling method. | 10 |
|  | Describe the measurement/rating process (e.g. time interval between repeated measurements, availability of clinical information, blinding). | 8 - 9 |
|  | State whether measurements/ratings were conducted independently. | 9 - 10 |
|  | Describe the statistical analysis. | 11 - 13 |
| RESUL TS | State the actual number of raters and subjects/objects which were included and the number of replicate observations which were conducted. | 13 |
|  | Describe the sample characteristics of raters and subjects (e.g. training, experience). | 13, Table 1 |
|  | Report estimates of reliability and agreement including measures of statistical uncertainty. | 13 - 16 |
| DISCUSSION | Discuss the practical relevance of results. | 17 - 20 |
| AUXILIARY MA TERIAL | Provide detailed results if possible (e.g. online) | Supplementary Materials |
