## Supplementary material for "Postoperative Determination of Directional Deep Brain Stimulation (DBS) Lead Orientation Using Rotational Fluoroscopy: Interobserver Agreement and Comparison with CT-Based Software": Online Appendix

### Circular Bland-Altman Analysis

#### Appendix

This document describes the mathematical formulation to calculate bias, limits of agreement (LoA), and confidence intervals in a Bland-Altman analysis for circular data (e.g. angles in radians or degrees). The following formula is used in the paper titled "Postoperative Determination of Directional DBS Leads Orientation Using Rotational Fluoroscopy: Interobserver Agreement and Comparision with CT-Based Softeware."

#### Given

Let $\theta_1, \theta_2 \in [-\pi, \pi]$ be the angular measurements (in radians) of two observers in the observations $n$.

#### 1. Circular Mean per observation

The circular mean for each pair of observations is given by:

$$\bar{\theta} = \arg\left(\frac{1}{2}\left(e^{i\theta_1} + e^{i\theta_2}\right)\right)$$

#### 2. Circular Differences

The signed circular difference is:

$$\Delta\theta = \text{circ\_dist}(\theta_1, \theta_2) = \text{angle}(e^{i(\theta_1 - \theta_2)})$$

#### 3. Bias and Standard Deviation

The mean bias (average difference) is:

$$\text{Bias} = \bar{\Delta\theta} = \frac{1}{n}\sum_{i=1}^{n} \Delta\theta_i$$

The (linear) standard deviation of the circular differences is:

$$\text{SD} = \sqrt{\frac{1}{n-1}\sum_{i=1}^{n}\left(\Delta\theta_i - \bar{\Delta\theta}\right)^2}$$

#### 4. Limits of Agreement (LoA)

The 95% limits of agreement are calculated as:

$$\text{LoA} = \text{Bias} \pm 1.96 \cdot \text{SD}$$

#### 5. Confidence Intervals (CI)

Let $t_{n-1,\alpha/2}$ be the $t$-value for a two-sided 95% confidence interval with $n - 1$ degrees of freedom.

##### Bias CI

$$\text{CI}_{\text{Bias}} = \text{Bias} \pm t_{n-1,\alpha/2} \cdot \frac{\text{SD}}{\sqrt{n}}$$

##### LoA CI

Following Bland and Altman, the standard error for LoA is:

$$\text{SE}_{\text{LoA}} = \sqrt{\frac{3 \cdot \text{SD}^2}{n}}$$

$$\text{CI}_{\text{LoA}} = \text{LoA} \pm t_{n-1,\alpha/2} \cdot \text{SE}_{\text{LoA}}$$
