## Supplementary Data for "Postoperative Determination of Directional Deep Brain Stimulation (DBS) Lead Orientation Using Rotational Fluoroscopy: Interobserver Agreement and Comparison with CT-Based Software"

**Angle Measurement by Rotational Fluoroscopy**

The Rotational Fluoroscopy is performed at the Cath lab in the Radiology Department at Jaslok Hospital, Mumbai, 24 hours after lead implantation. The patients were lying supine in the Cath lab with their heads facing upwards. Standard anteroposterior (AP) and lateral x-rays were obtained first, ensuring that the collimation beam was aligned parallel to the orbito-meatal and bi-meatal axes, respectively. Subsequently, a 210°–240° rotational fluoroscopy was performed using a flat-panel C-arm system. The system acquired ~120–200 frames during a 4–5 second sweep. Radiation exposure (dose-area product ~2.3 mGy·cm²) is comparable to four plain skull x-rays and substantially lower than a conventional head CT (effective dose ~0.2 mSv vs. ~2.3 mSv).

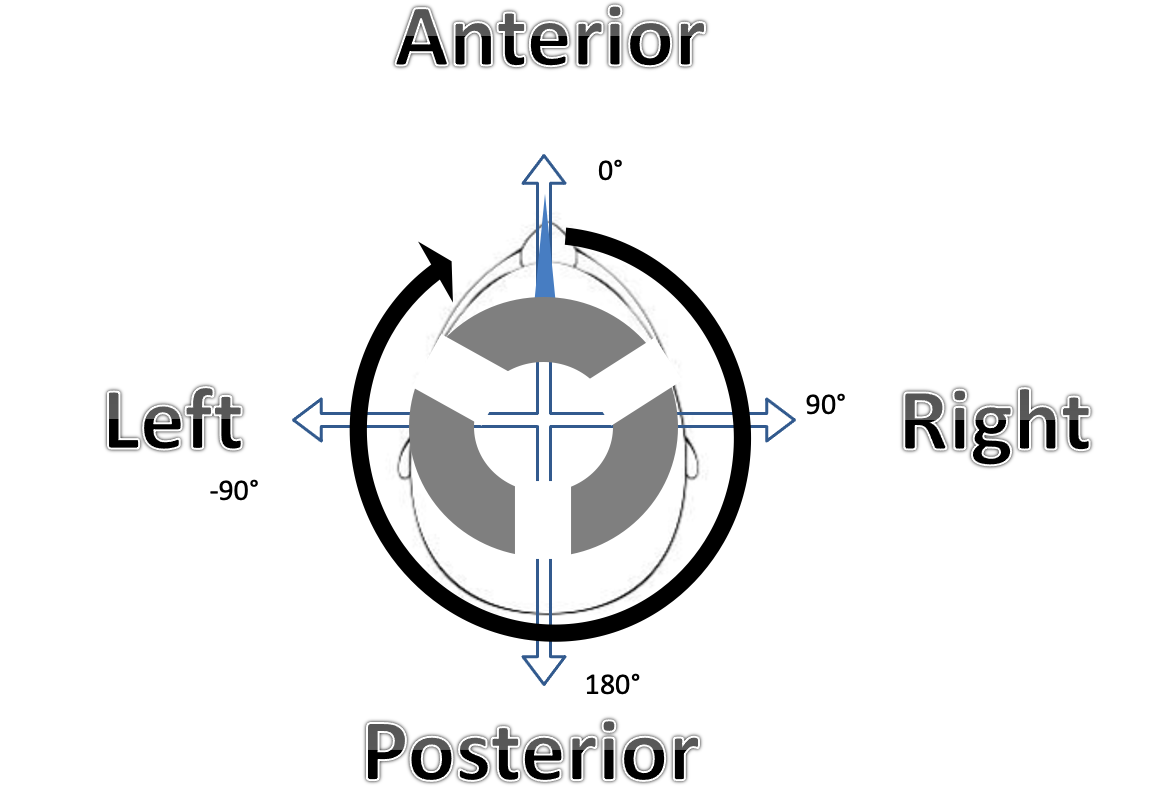

As shown in the Figure, the anterior skull midline was defined as 0°. Clockwise rotations were assigned positive values; counterclockwise rotations were negative. AP and lateral views were used to determine the gross quadrant of the x-ray marker (anterior, posterior, right, or left). Modern directional leads have segmented distal contacts separated by narrow gaps. As the fluoroscopy rotates around the patient’s head, these gaps align at specific projection angles (30°, 90°, 150°, 210°, 270°, and 330°). When alignment occurs, the gaps form a distinct straight radiolucent line resembling the “iron sights” of a firearm. This pattern provides a reproducible visual cue to the lead’s rotational orientation.

Observers scroll through the reconstructed rotational dataset to identify the projections where the segment gaps overlap into an iron sight. The detector angle at which this occurs is recorded. Final orientation is calculated by subtracting the C-arm collimation angle from the observed “iron sight” angle. as mentioned by Reinacher et al^1^.
