## Supplementary Tables for "Postoperative Determination of Directional Deep Brain Stimulation (DBS) Lead Orientation Using Rotational Fluoroscopy: Interobserver Agreement and Comparison with CT-Based Software"

Table A. Imaging Acquisition Parameters for Postoperative CT and Rotational Fluoroscopy in DBS Patients

| Modality | Parameters |
| --- | --- |
| CT | 64-section Dual energy CT scanner (Discovery 750 GE healthcare), 1mm pitch, x 1 mm slice thickness field of view 300 mm, 250 slices, tilt 0°, helical mode, voltage 140 kV, exposure 500 mAs |
| Fluoroscopy | Philips Allura Xper FD20, motorized automated rotational scan, Matrix 2048 x 2048, pixel size 12-bit, Source to Image (SID) distance 120 cm, using voltage 80 kV, 260 mAs and 30 frames per second without any digital subtraction |

Imaging acquisition parameters for postoperative CT and rotational fluoroscopy used to determine directional DBS lead orientation in patients at Jaslok Hospital, Mumbai, between 2022 and 2024. CT scans were processed via Brainlab Elements™, and 3D rotational fluoroscopy images were analyzed using the Iron Sight method.

Table B. Calculation Parameters for Interobserver Agreement Using Iron Sight Method

| SS_total | SS_subject | SS_rater | SS_error | MS_subject | MS_rater | MS_error | df_subject | df_rater | df_error |
| --- | --- | --- | --- | --- | --- | --- | --- | --- | --- |
| 12.34688 | 12.23776 | 0.004971 | 0.104149 | 0.177359 | 0.004971 | 0.001509 | 69 | 1 | 69 |

Components of the Intraclass Correlation Coefficient (ICC (2,1)) calculation for interobserver agreement in directional DBS lead orientation using the Iron Sight method on postoperative rotational fluoroscopy. Data from 70 leads implanted between 2022 and 2024 at Jaslok Hospital, Mumbai. SS- Sum of Squares, MS- Mean Square, df- degree of freedom

Table C. Calculation Parameters for Agreement Between Imaging Modalities

| SS_total | SS_subject | SS_within | SS_error | MS_subject | MS_error | df_subject | df_rater | df_error |
| --- | --- | --- | --- | --- | --- | --- | --- | --- |
| 7.615354 | 7.094762 | 0.520591 | 0.520591 | 0.107496 | 0.007888 | 66 | 1 | 66 |

Components of the Intraclass Correlation Coefficient (ICC (3,1)) calculation for method agreement between Iron Sight-derived and Brainlab Elements™ CT-derived orientation estimates of directional DBS leads. Analysis includes 67 leads from patients treated at Jaslok Hospital, Mumbai, between 2022 and 2024. SS- Sum of Squares, MS- Mean Square, df- degree of freedom
